# Determination of utility of 3 minute and 6 minute walk tests in assessment of progression of mild COVID-19 infection at a tertiary hospital in North India

**DOI:** 10.1101/2021.10.03.21264476

**Authors:** Aastha Goel, Souradeep Chowdhury, Tamoghna Ghosh, Anupam Singh, Arvind Kumar, Naveet Wig

## Abstract

**Introduction:** There have been 214 million confirmed cases of COVID-19 worldwide with a total death tally of 4.4 million. The current study aims to determine the predictive value of 3 minute and 6-minute walk tests in assessment of progression of mild COVID-19 infection at a tertiary care hospital in North India.

**Methods:** The study population consisted of adults (age more than 18 years) with a confirmed diagnosis of Covid-19 by RT-PCR on nasopharyngeal specimens. Patients with only mild illness were enrolled. After the patients were admitted to the isolation ward, the presenting history, comorbidity status, vital signs and laboratory parameters were recorded. The 3 and 6 minute walk test was performed daily from admission till discharge or progression of severity of COVID-19 and it was used to calculate BDS and NEWS2 scores.

**Results:** Our study consisted of 50 patients with 34 (68%) males and the mean (SD) age of the patient population being 28.1 (6.4) years. The most common symptoms were fever, sore throat, and cough. All laboratory parameters were within normal ranges for all the patients. 96% recovered without progression, while only 4% of them progressed to moderate illness. Results of the 3 and 6 minutes walk tests, BDS and NEWS2 scores showed improvement over the course of hospital stay.

**Conclusions:** Although the walk tests and the scores improved over time, they failed to predict the disease progression.

## Introduction

The ongoing pandemic of coronavirus disease (Covid-19) is caused by Severe Acute Respiratory Syndrome coronavirus 2 (SARS-CoV-2). It has posed a serious threat to public health worldwide. As of 25th August 2021, there have been 214 million confirmed cases of covid-19 worldwide with a total death tally of 4.4 million.^1^ Covid-19 can be categorised into asymptomatic, mild, moderate, severe and critical illness based on respiratory rate and blood oxygen saturation. These categories may overlap or the patients’ clinical status may change with time.^2^

Despite being into a year of the pandemic, no antiviral agent has been found to be effective. Although vaccines have rolled out with warp speed, recent outbreaks with mutant strains of the virus have put their efficacy into question.^3,4^ Numerous factors have been associated with increased risk of severe COVID-19 infection. It includes diabetes, old age, lower lymphocyte count and a higher LDH values.^5,6^ A large majority of patients (80%) have mild infection with an uneventful recovery. Mild infection has the potential of rapidly worsening into severe Covid-19 infection with respiratory failure. Therefore, there is a need to recognize the patients with potential for deterioration early in the course of illness.

The current study aims to determine the predictive value of 3 minute and 6-minute walk tests in assessment of progression of mild COVID-19 infection at a tertiary care hospital in North India. This would help in developing a triage system of patients with mild COVID-19 with high likelihood of deterioration so that early steps can be taken for referral.

## Methods

### Study population

The study was an ambispective, analytical, observational study conducted at a tertiary care teaching hospital in New Delhi, India. The study population consisted of adults (age more than 18 years) with a confirmed diagnosis of Covid-19 by RT-PCR on nasopharyngeal specimens. Patients with only mild illness were enrolled. Mild illness is defined as a room air saturation of more than 94% and a respiratory rate of less than or equal to 24. Progression to moderate/severe COVID-19 was defined as respiratory rate more than 24 or room air saturation of less than or equal to 94% or new onset oxygen requirement. In view of the infectious nature of the disease telephonic consent was obtained. The study was approved by the Ethics Committee of All India Institute of Medical Sciences, New Delhi, India.

### Data Collection

A formal sample size calculation was not done and instead convenience sampling was used. After the patients were admitted to the isolation ward, the presenting history, comorbidity status and vital signs were recorded. The laboratory parameters, including complete blood count, liver and renal function test, lactate dehydrogenase (LDH), C-reactive protein (CRP), ferritin, erythrocyte sedimentation rate (ESR) and coagulation profile were obtained at admission. A baseline plain chest radiograph and ECG were also obtained at admission.

### Patient Performance Measurement

The 6 minute walk test was performed daily from admission till discharge or progression of severity of COVID-19. It was performed in the corridor of the isolation ward. The distance between the two ends of the corridor was measured and estimated to be 25 meters. It was marked for convenience. The patients were asked to walk between the two ends and the distance measured covered and the oxygen saturation at baseline, 3 minute and 6 minute was recorded. The data that was obtained was also used to calculate the Borg dyspnea score (BDS) and the National Early Warning Score 2 (NEWS2). Developed by Gunnar Borg, the score allows individuals to subjectively rate their level of exertion during exercise or exercise testing (American College of Sports Medicine, 2010).^7^ The National Early Warning Score (NEWS) was launched by the Royal College of Physicians (RCP) in 2012 to improve the identification, monitoring and management of unwell patients in hospitals.^8^ It is based on a logistic regression model designed to predict in-hospital patient mortality within 24 hours of a set of vital sign observations.^9^ Originally consisting of pulse rate, respiratory rate, blood pressure, temperature and oxygen saturation, it was updated in 2017 to NEWS2, which incorporated a new onset of confusion and a separate scoring system for oxygen saturation in patients with type 2 respiratory failure.

### Statistical Analysis

The association between the categorical variables was statistically analyzed by the chi-square test. Difference of means of 2 groups was calculated by t test. Statistical analysis was performed using StataCorp. 2015. Stata Statistical Software: Release 14. College Station, TX: StataCorp LP, and a p< 0.05 was considered statistically significant.

## Results

A total of 50 patients, with mild COVID-19 disease at the time of presentation, were screened for inclusion in the study. Among them 34 (68%) were males and the mean (SD) age of the patient population was 28.1 (6.4) years. The most common symptoms were fever (42; 84%), sore throat (33; 66%), cough (27; 54%), loss of smell (24; 48%), loss of taste (23; 46%), rhinorrhoea (20; 40%), loose stool (7; 14%), and dyspnoea (4; 8%). None of the patients had chest X ray abnormalities, while ECG was normal in 49 (98%) of the patients. Hematological and inflammatory parameters like NLR (Neutrophil Lymphocyte Ratio), Ferritin, LDH, C-reactive Protein and D-dimer were within normal ranges for all the patients at the time of testing on admission, owing to the mild nature of the disease. 34 (68%) of the patients didn’t have any comorbidities, while among those who had comorbidities (32%), hypertension, diabetes and obstructive airway diseases were the most common ones. Among the 50 mild illness patients, 48 (96%) recovered without progression, while only 2 (4%) of them progressed to moderate illness. The mean (SD) time of recovery was 8 (4.9) days.

Among the predefined scales that were used to follow up the patients of mild COVID, the best predictor was found to be Borg dyspnea scale. The mean score was 0.01 on Day 1 and there was no statistically significant difference between the Day 1 score and any subsequent day score (Days 2-10). (figure 1) In the NEWS2 scoring, the mean score decreased from Day 5 onwards and all of them were significantly lower than Day 1 mean score. (figure 2) In either of the 3 and 6 Minutes Walk Test, mean SpO2 on Day 1 was not statistically different from any subsequent day (Days 2-10). (figure 3) In the mean distance covered in the 6 Minute Walk Test, on 7 out of 9 days, following Day 1, the mean distance was statistically greater than the mean of Day 1. (figure 4)

**Figure 1:**
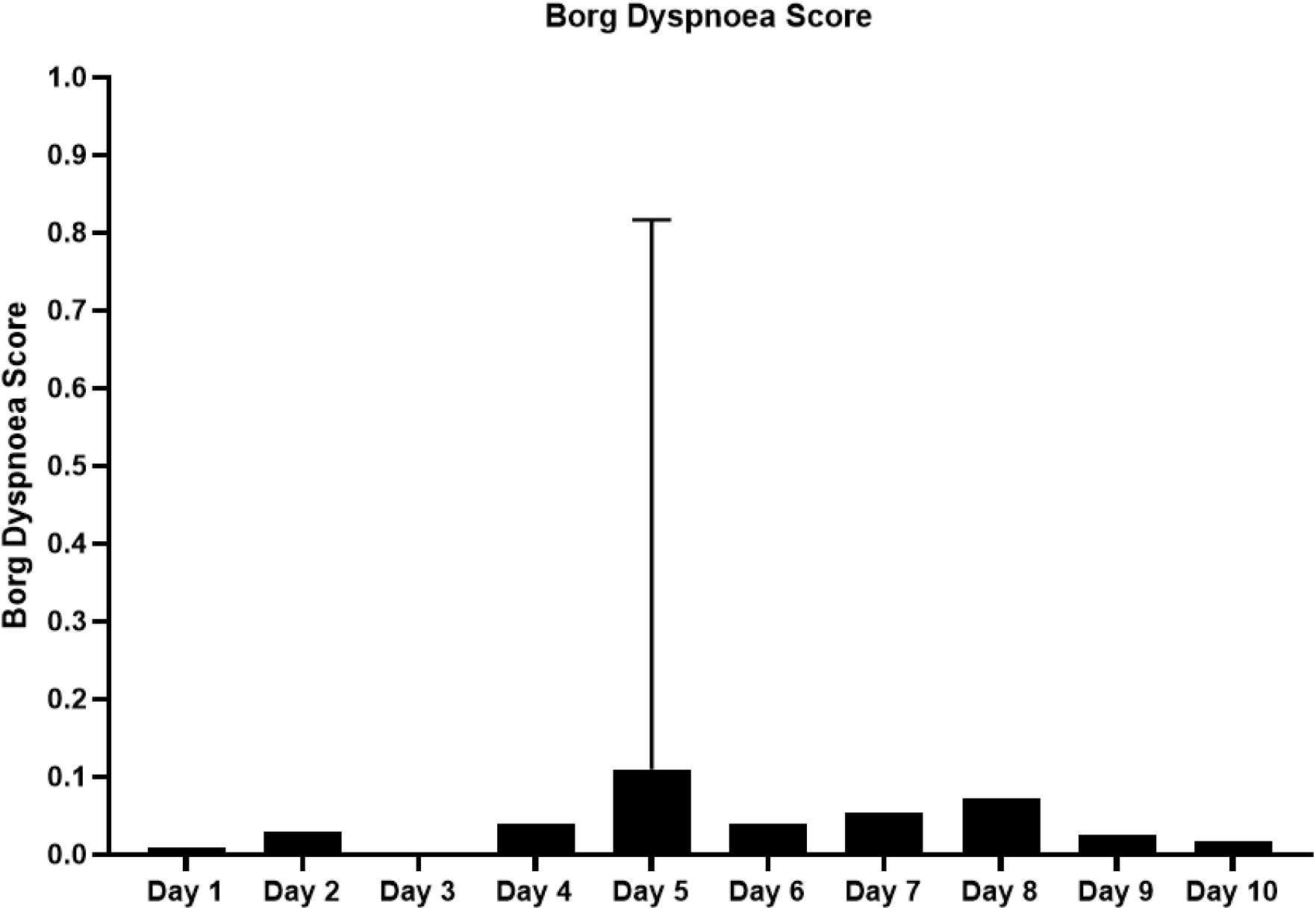
Mean (SD) of Borg Dyspnoea Score from Day 1 to Day 10 of hospital admission. There was no statistically significant difference between the Day 1 score and any subsequent day score (Days 2-10).

**Figure 2:**
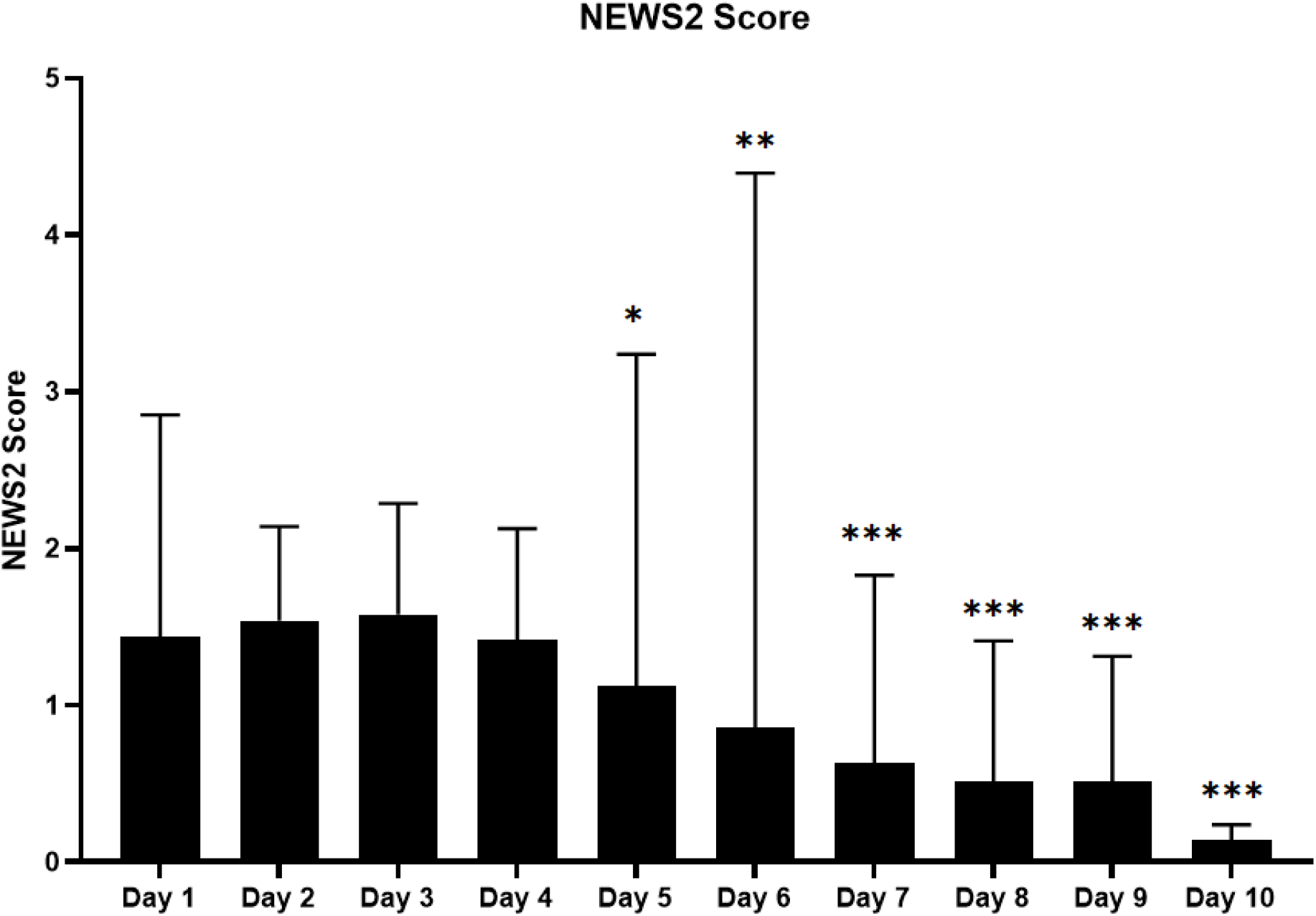
Mean (SD) of NEWS2 score from Day 1 to Day 10 of hospital admission. The mean score decreased from Day 5 onwards and all of them were significantly lower when compared with Day 1 mean score. Here *means p<0.05, ** means p<0.01, and *** means p<0.001

**Figure 3:**
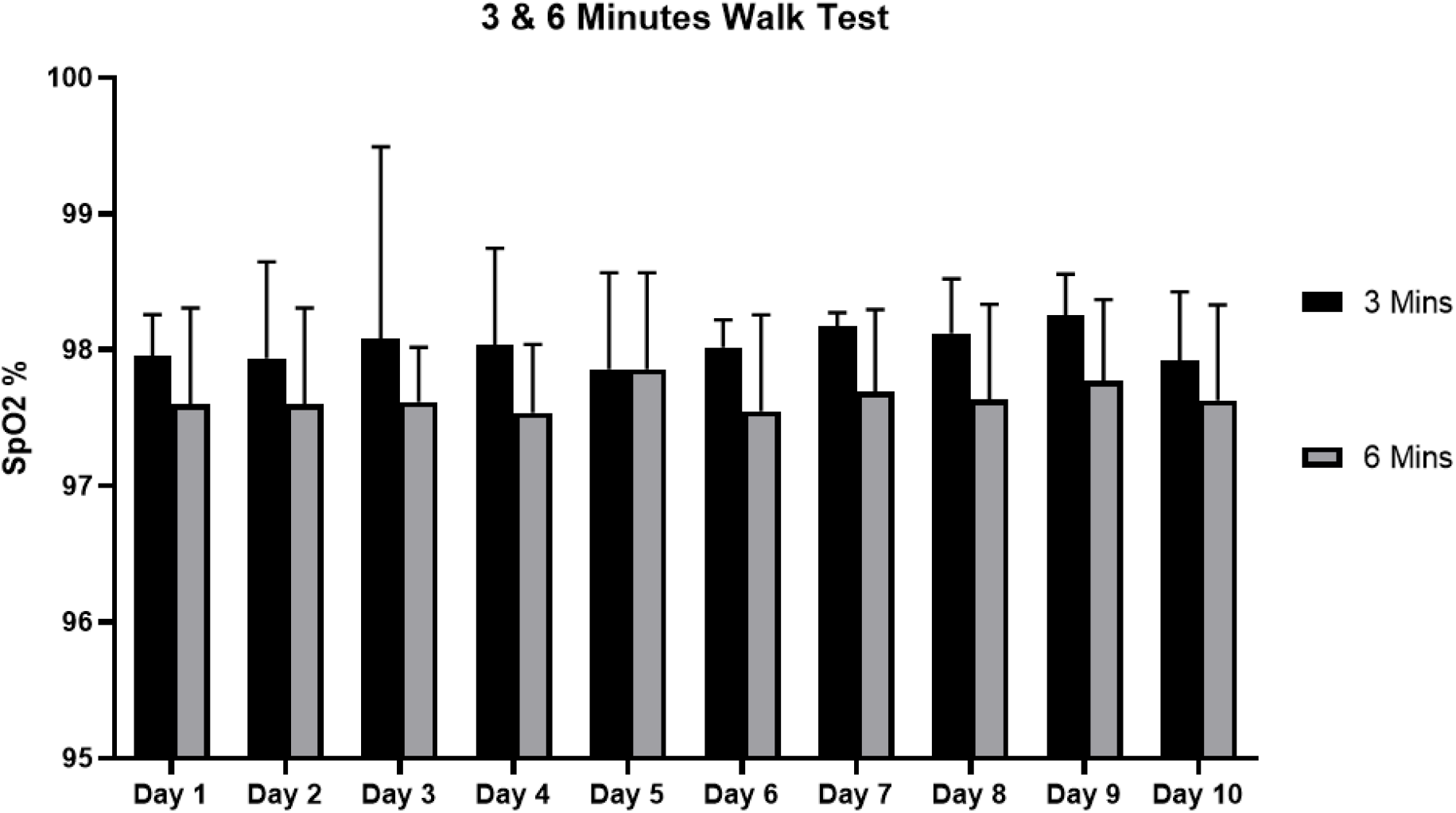
Mean (SD) of SpO_2_ in 3 and 6 Minutes Walk Tests from Day 1 to Day 10 of hospital admission. In either of the 3 and 6 Minutes Walk Test, mean SpO_2_ on Day 1 was not statistically different from any subsequent day (Days 2-10).

**Figure 4:**
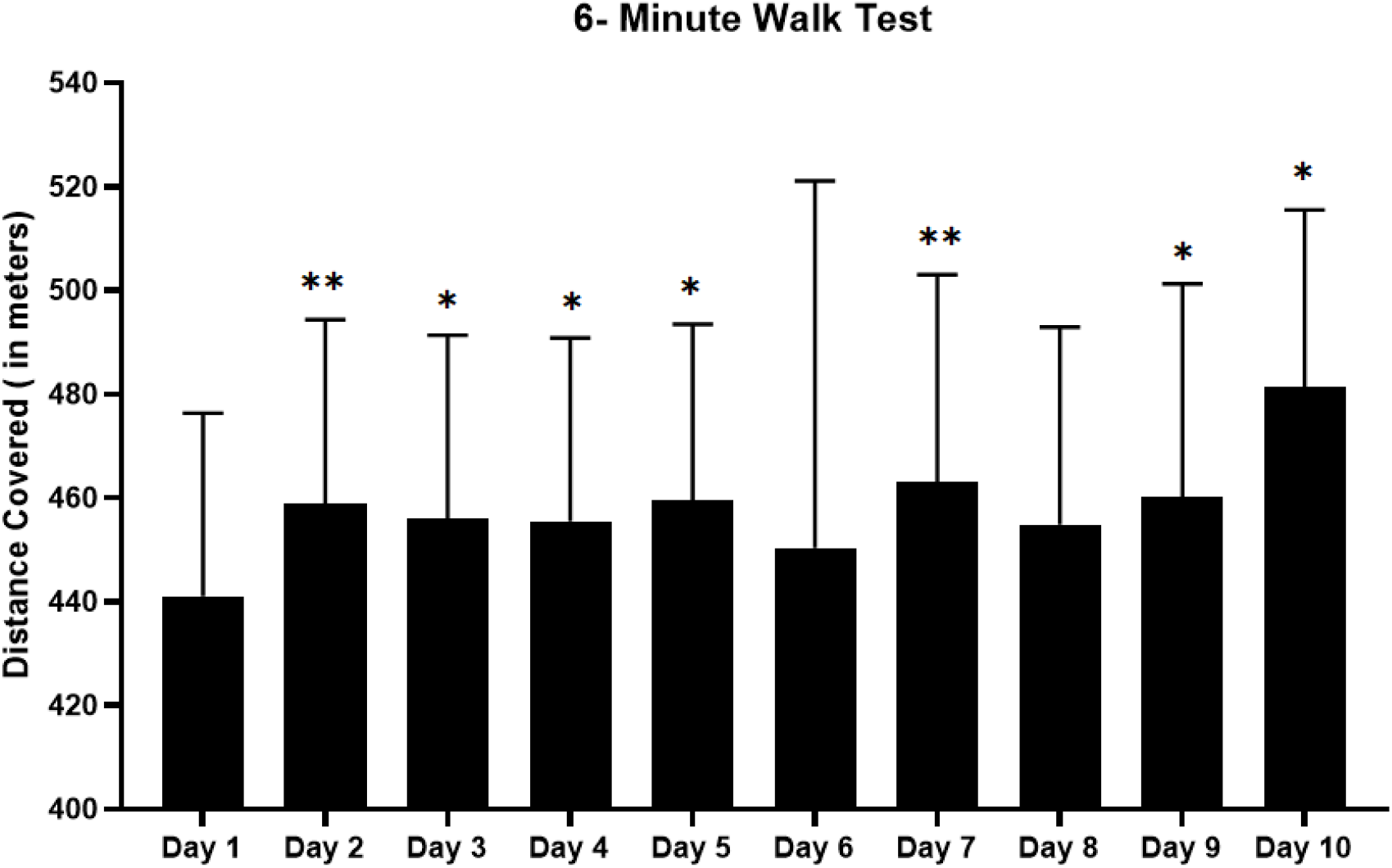
Mean (SD) of distance covered (in meters) in 6 Minutes Walk Tests from Day 1 to Day 10 of hospital admission. On Days 2,3,4,5,7,9, and 10, the mean distance was statistically greater than that of Day 1. Here *means p<0.05, and ** means p<0.01

Among the other parameters assessed in the study, only the Borg dyspnea scale, assessed at Day 2 of admission, predicted time to recovery, and it was statistically significant. The use of the Borg dyspnea scale early on in the disease process would help to identify those patients who might be predicted to have a longer course, and that would help them to seek medical help and hospitalisation early.

## Discussion

Various scoring systems have been devised to predict the progression of COVID-19 in patients, and a recent systematic review has shown that as many as 30 risk factors could be helpful in the development of a prediction tool.^10^ Of these, the NEWS score has been validated for predicting severe disease, and has a sensitivity and specificity of 80% and 83.4% respectively.^11^

Traditionally, the 6-minute walk test has been used to determine the cardiopulmonary reserves and as a one-time measure of functional status of patients, as well as a predictor of morbidity and mortality in moderate to severe heart and lung disease.^12^

In a time when we were scurrying to repurpose the therapeutic agents to combat the rapid influx of COVID-19 patients, it seemed only prudent to explore other scoring systems and predictive models to better stratify such patients. This would be of particular help at a primary care level, where patients predicted to worsen could be better triaged and referred to higher centres for quick medical attention. Also, once the government had approved home isolation as an accepted alternative, patients themselves could be trained to seek help, based on scoring systems that are easy to administer.

So we attempted to assess the utility of the 6 minute and 3 minute walk tests in the context of COVID-19. In our patient population, only two patients progressed to moderate COVID-19, on day 7 and day 10 of their illness. In both these patients, the 6 minute and 3 minute walk tests failed to predict the progression of disease. Also, the validated NEWS score for both patients was 5, which falls in the medium risk category. So the practice of enthusiastically determining the NEWS2 score in all mild COVID patients, with the understanding that it accurately classifies the stage of the disease, may be unfounded. Coming to the results of the 6 minute and 3 minute walk test, both the distance covered as well as the difference in SpO2 failed to provide statistically significant data, to predict the recovery or progression of our subset of patients. Since our patient cohort was small, further follow up studies are needed to formalise the use of these scores, and their subsequent real world utility.

The patients were discharged as per hospital policy on day 10 of illness, and were followed up telephonically to assess resolution of their symptoms. This helped us assess the various calculated parameters and their utility in predicting the time to recovery.

Further studies are needed on this, with larger sample sizes, to determine the usefulness of this scale. Further data would put this on a firm footing and help us in using easily administered scales to triage patients and stratify them based on risk.

## Data Availability

Data will be made available on request

## Acknowledgements

The authors express their sincere gratitude to the Department of Medicine for sample collection, and data entry staff and technicians of the Department of Medicine, All India Institute of Medical Sciences, New Delhi, India for their assistance in procuring the clinical and laboratory details and preparing the raw data.

## Funding

This was an investigator-initiated non-funded study

## Availability of data and materials

The data would be made available by the authors on specific request keeping patient confidentiality in view.

## Competing interests

The authors declare that they have no competing interests.

## Notes

### Competing Interest Statement

The authors have declared no competing interest.

### Funding Statement

This study received no external funding

### Author Declarations

Institute Ethics Committee All India Institute of Medical Sciences

